# SARS-CoV-2 NSP3, NSP4 and NSP6 mutations and Epistasis during the pandemic in the world: Evolutionary Trends and Natural Selections in Six Continents

**DOI:** 10.1101/2022.05.22.22275422

**Authors:** Haniyeh fooladinezhad, Maryamsadat Shahidi, Mohammadamin Mahmanzar, Bahar Mahdavi, Samaneh Tokhanbigli, Mahsa Mollapour Sisakht, Bahman Moradi, Mohamad Reza Ganjalikhany, Karim Rahimian, Mazdak Ganjalikhani-Hakemi

## Abstract

**Background:** The Coronavirus 2019 (COVID-19) was named by the World Health Organization (WHO) due to its rapid transmittable potential and high mortality rate. Based on the critical role of None Structural Proteins (NSP), NSP3, NSP4, and NSP6 in COVID-19, this study attempts to investigate the superior natural selection mutations and Epistasis among these none structural proteins.

**Methods:** Approximately 6.5 million SARS-CoV-2 protein sequences of each NSP3, NSP4, and NSP6 nonstructural protein were analyzed from January 2020 to January 2022. Python programming language was utilized to preprocess and apply inclusion criteria on the FASTA file to prepare a list of suitable samples. NSP3, NSP4, and NSP6 were aligned to the reference sequence to compare and identify mutation patterns categorized based on frequency, geographical zone distribution, and date. To discover epistasis situations, linear regression between mutation frequency and date among candidate genes was performed to determine correlations.

**Results:** The rate of NSP3, NSP4, and NSP6 mutations in divided geographical areas was different. Based on continental studies, P1228L (54.48%), P1469S (54.41%), and A488S (53.86%) mutations in NSP3, T492I (54.84%), and V167L (52.81%) in NSP4 and T77A (69.85%) mutation in NSP6 increased over time, especially in recent months. For NSP3, Europe had the highest P1228L, P1469S, and A488S mutations. For NSP4, Oceania had the highest T492I and V167L mutations, and for NSP6, Europe had the highest T77A mutation. Hot spot regions for NSP3, NSP4, and NSP6 were 1358 to 1552 AA, 150 to 200 AA, and 58 to 87 AA, respectively. Our results showed a significant correlation and co-occurrence between NSP3, NSP4, and NSP6 mutations.

**Conclusion:** We conclude that the effect of mutations on virus stability and replication can be predicted by examining the amino acid changes of P1228L, P1469S, A488S, T492I, V167L and T77A mutations. Also, these mutations can possibly be effective on the function of proteins and their targets in the host cell.

## 1. Introduction

At the end of 2019, a virus from the Coronavirus family (nCoV-2019) became the major cause of spreading infectious lung disease [1]. It presented by the International Committee on Taxonomy of Viruses (ICTV) as the Severe Acute Respiratory Virus 2 (SARS-CoV-2) and named the Coronavirus 2019 (COVID-19) via the World Health Organization (WHO) [2]. In a short time, the epidemy became a global outbreak that affected all countries [3]. The origin of this virus has not yet been determined. However, bioinformatics analyses of viral isolates in different countries have different origins for that [4]. The highest mortality rate of Covid-19 disease, so far, has been related to people with a history of underlying diseases such as hypertension, diabetes, cancer, cardiovascular disease, or chronic respiratory disease [5, 6].

SARS-CoV-2 is a unipolar RNA virus, a positively polarized strand with a genome size of 29903 nucleotides. The virus genome consists of two untranslated regions (UTRs) at the 5□ and 3□ ends and eleven open reading frameworks (ORFs) that encode 27 proteins [7, 8]. The first ORF (ORF1 / ab) makes up about two-thirds of the virus genome and encodes sixteen non-structured proteins (NSPs) [9]. The remaining one-third of the genome encodes four structural proteins and at least six ancillary proteins. Structural proteins are: spike (S), small envelope (E), matrix (M), and phosphor-protein nucleocapsid (N), and side proteins are: ORF3a, ORF8, ORF7a, ORF7b, ORF10, and ORF6 [10-13] (Fig 1A). 5′-UTR and 3′-UTR of COVID-19 include 265 and 229 nucleotides, respectively. ORF1ab (GeneID: 43740578) contains 21,290 nucleotides. It encodes the pp1a protein containing 4405 amino acids (NSP1-NSP11) or the pp1ab protein containing 7096 amino acids (NSP1-NSP16) by using a ribosomal shift frame (Fig 1B). The NSP1 protein suppresses the antiviral response of the host [14], NSP3 is a papain-like protease [15, 16], NSP5 is a 3CLpro (the second 3C-like protease) [17], NSP7 and NSP8 form a complex to create a primase [18], NSP9 can bind RNA/DNA [13], NSP12 is RNA polymerase dependent to RNA (Rdrp) [19], NSP13 is a helicase [20], NSP14 is exonuclease 3□ to 5□ (Exon) (6), and NSP15 is a specific poly U endoribonuclease (XendoU) [21]. The extant NSPs are played a rule in the transcription and replication of the viral genome [12, 22]. NSP3, NSP4, and NSP6 are transmembrane proteins that have roles in regulating host immunity, and improving the function of host cell organelles for viral replication, and selected as study purposes [23]. In between, NSP3 is a key component and the largest NSP involved in the replication and transcription of the virus. It is located on the host cell membrane, where the replication and transcription of the virus genome occur [24-26]. NSP3 is a multidomain protein that contains an N-terminal Ubl (ubiquitin-like domain), followed by an ADRP (ADP-ribose phosphatase), SUD (SARS-unique domain) domain, and another Ubl domain which is a part of PLpro. C-terminal of the protease is a NAB (nucleic acid binding) domain followed by a T.M. (transmembrane segment), ZnF (zinc finger) domain, and also α -helical regions that may be effective in connecting NSP3 to NSP4. All of these domains place in host cell cytoplasm [27, 28]. One of the important domains involved in transcription is SUD, which interacts with guanine-quadruplex (G4) structures. Whereas, G4s are non-canonical nucleic acid structures organized by virus G-rich RNA or DNA sequences and supervise critical points of viral replication. It plays a main role in forming the replication/transcription viral complex (RTC) [29, 30]. Nevertheless, some of the functions of NSP3 are still unknown [31]. NSP4 with transmembrane domain 2 (TM2), a transmembrane glycoprotein, is better defined, participates specially in the formation of the double-membrane vesicles (DMV), and have copies of virus fragments that associated with replication complexes [32, 33]. NSP6 protein with putative transmembrane domain blocks ER-induced autophagosome/autolysosome vesicle formation and plays a protective function in checking viral production in host cells. NSP6 with T.M. domain, via the activation of the omegasome pathway, enforces autophagy which has acted in favor of causing infection by the virus [34] (Fig 1C). These proteins (NSP3, NSP4, and NSP6) are predicted to be integral membrane proteins. They probably have a main role in anchoring the replication complexes into the lipid bilayer [32, 33].

**Fig. 1.**
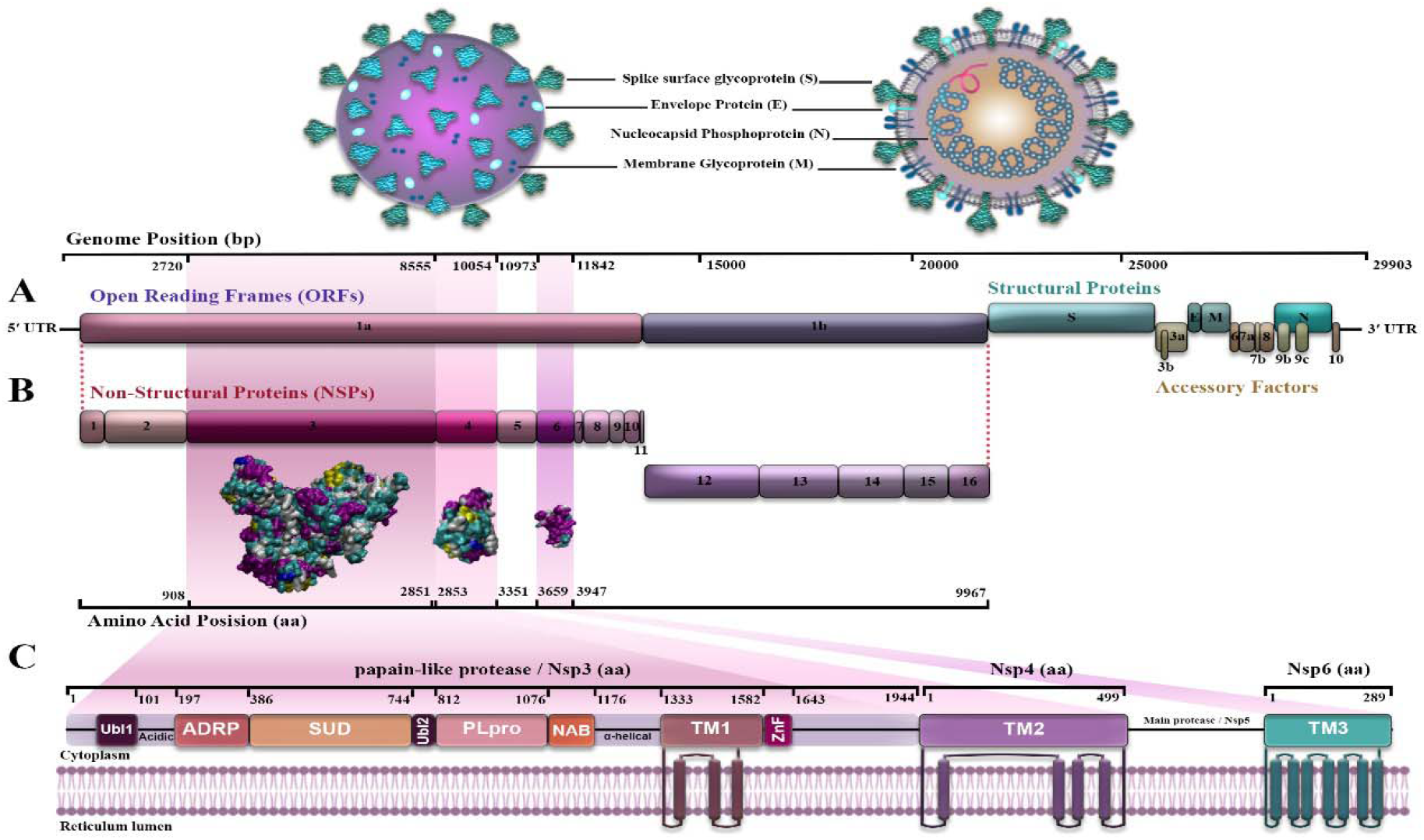
Structure and genome of acute severe respiratory coronavirus syndrome-2 (SARS CoV-2) (8, 9, 10, 11); A) The four structural proteins are as follows: spike (S), envelope (E), membrane (M), nucleocapsid (N); B) Location of non-structured proteins: NSP3, NSP4 and NSP6 from 5′-UTR to 3′UTR on the virus genome. C) NSP 3, NSP4 and NSP6 positions and domains.

In this context, the most important issue is to investigate the key mutations in nonstructural proteins NSP3, NSP4, and NSP6, obtained from patients with SARS-CoV-2 worldwide. Our goal was to predict the effect of amino acid changes based on their hydrophobicity (HΦ) according to previous studies on protein stability and replication. Clustering results led us to investigate high score mutions that could effects on hydrophobic residues and accessible to potential immune responibilties. It may aid to the development of new strategies for the treatment and prevention of COVID-19.

## 2. Materials and Methods

### 2.1 Sequence source

This study fully evaluated relevant data about the amino acid (AA) sequences (AAS) of NSP3, NSP4 and NSP6 in SARS-CoV-2. The sequence of all amino acids was compared with the AAS reference sample of the Wuhan-2019 virus with access number “EPI_ISL_402124”. All data were retrieved from January 2020 until January 2022 from GISAID (www.gisaid.org) [35-37]. Database through coordination with Erasmus Medical Center (Fig. 2A).

**Fig. 2.**
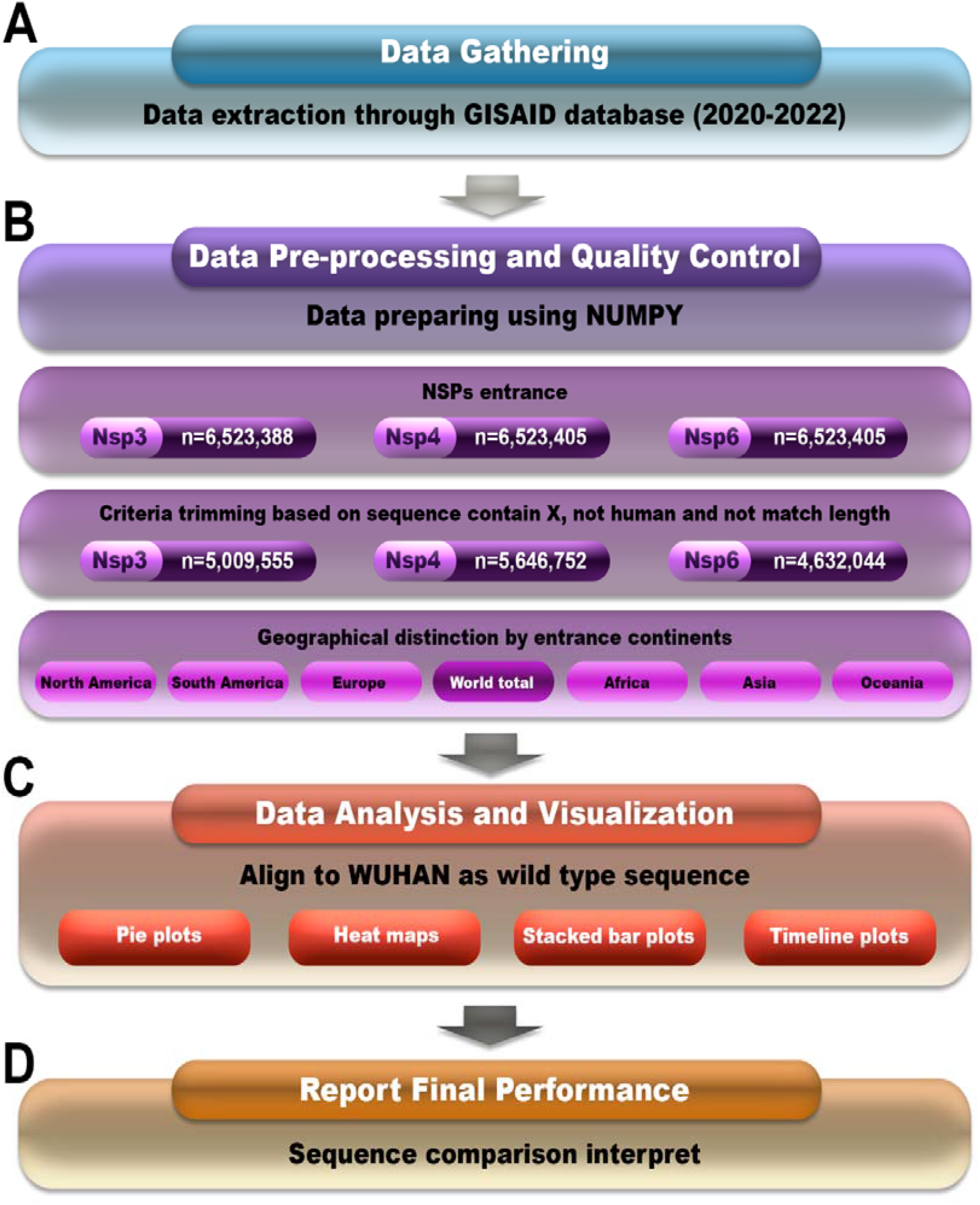
Data processing workflow used to demonstrate and validate NSP3, NSP4 and NSP6 AAs mutations. A) Data Gathering B) Data Pre-processing and Quality Control C) Data Analysis and Visualization D) Report Final Performance

### 2.2 Sequence analyses and processing

Python 3.8.0 language programming and ′Numpy′ and ′Pandas′ libraries were used in this research to preprocess FASTA files, extract NSP3, NSP4, and NSP6 from other genes, and perform sequence alignment. The NSP3 AAs are located between 2720 and 8554, NSP4 consists of AAs between 8555 and 10054, and NSP6 comprises AAs between 10973 and 11842, all of which are located on the ORF1ab sequence.

Every difference between sample and reference sequence was identified as a mutation, and the position and exchanged AA were reported. Non-human specimens (such as bats and pangolins), those with more or less than 126229 AA for NSP3, 12568 AA for NSP4, and 1477693 AA for NSP6, and specimens containing un-specified AAS (reported as X) were excluded. Finally, 5009555 samples for NSP3, 5646752 samples for NSP4, and 4632044 samples for NSP6 were included in this study.

The knowing algorithm for finding mutants was as follows:

As all sequences have equal lengths, the following suggested algorithm used ‘Refseq’ and ‘seq’ that point to reference sequence and sample sequence, respectively.

For refitem, seqitem in zip (refseq, seq)

If (refitem! =seqitem)

Report a new mutant

After retrieving NSP3, NSP4, and NSP6, each sample’s continent name and geographical coordinates were received and then mapped global outbreaks of mutations by pycountry-convert 0.5.8 software and ‘Titlecase’ library in Python. After that, The mutations were classified into six geographical regions: North America, South America, Europe, Asia, Oceania, and Africa (Fig. 2B).

Normalized frequency of each region was used to report data and draw graphs. To normalizing data, divided number of mutations by number of its continent sequences in R 4.0.3 programming envirment(Fig. 2C). Our analysis workflow and filterings are summarized in Fig.2.

### 2.3 Estimation of Epistasis from NSP3,4,6

In this study, assuming that we have only one variable, the number of mutations in each sample, we considered the relationship of these variables to be linearly and non-parametric.

F(x) = B1X+B0

Therefore, to modeling and evaluate distinct scaler relationships between NSP3, NSP4 and NSP6 by used Stat Package in phyton language programming environment, the linear regression model was predicted.

## 3. Results

### 3.1 Distribution of NSP3, NSP4, and NSP6 Mutations Per Continent

To evaluate the distribution of mutations in NSP3, NSP4, and NSP6, mutation evaluation analyses were performed on 5009555 AASs samples of NSP3 (1945 AA. length), 5646752 AASs samples of NSP4 (with 500 AA length), and AASs 4632044 samples of NSP6 (with 290 AA length). Data retrieved from GISAID database (www.gisaid.org) from January 2020 to January 2022 [35-37]. Supposed identified mutation frequency, NSP3, NSP4, and NSP6 aligned with the reference sequence.

The results appeared in table 1 show the number of NSP3, NSP4, and NSP6 mutations (Fig. 3). In NSP3, we observed that over 47% of our samples had more than four mutations, and had the lowest mutation rate among NSP3, NSP4, and NSP6 which indicates NSP3 is highly mutable.

**Table 1.**
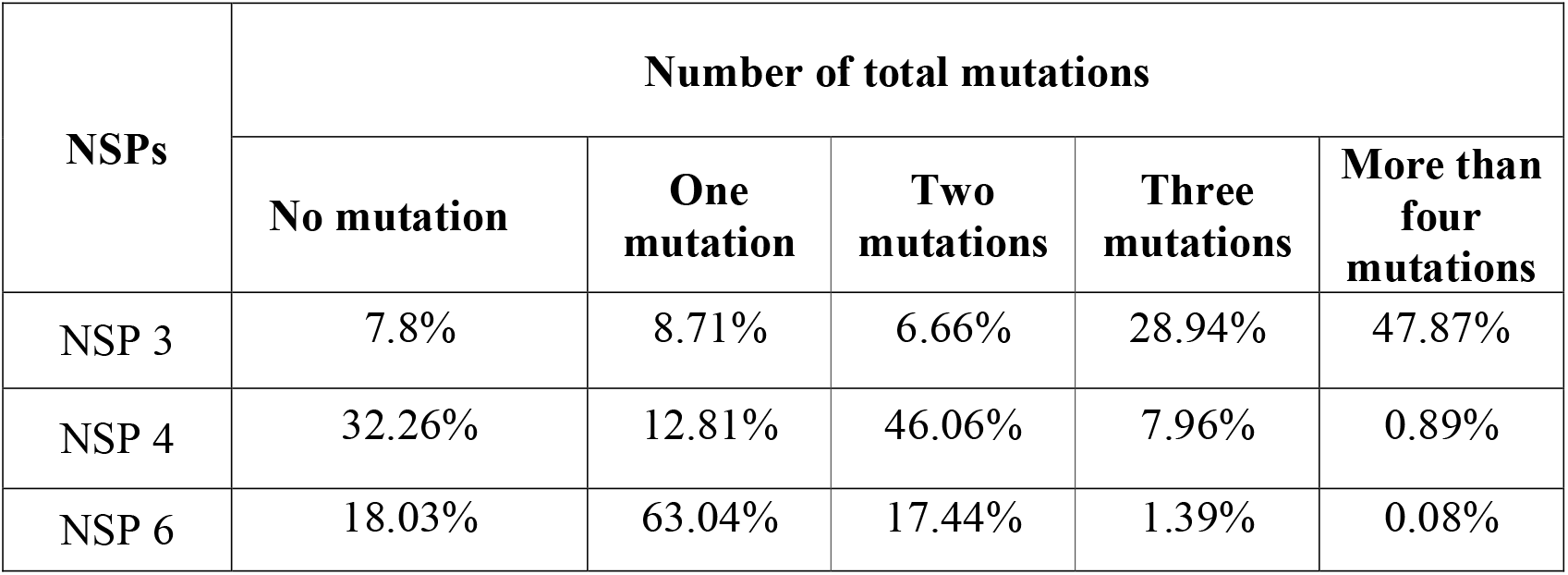
The number of total NSP 3, 4, and 6 mutations

| NSPs | Number of total mutations |  |  |  |  |
| --- | --- | --- | --- | --- | --- |
|  | No mutation | One mutation | Two mutations | Three mutations | More than four mutations |
| NSP 3 | 7.8% | 8.71% | 6.66% | 28.94% | 47.87% |
| NSP 4 | 32.26% | 12.81% | 46.06% | 7.96% | 0.89% |
| NSP 6 | 18.03% | 63.04% | 17.44% | 1.39% | 0.08% |

**Fig. 3.**
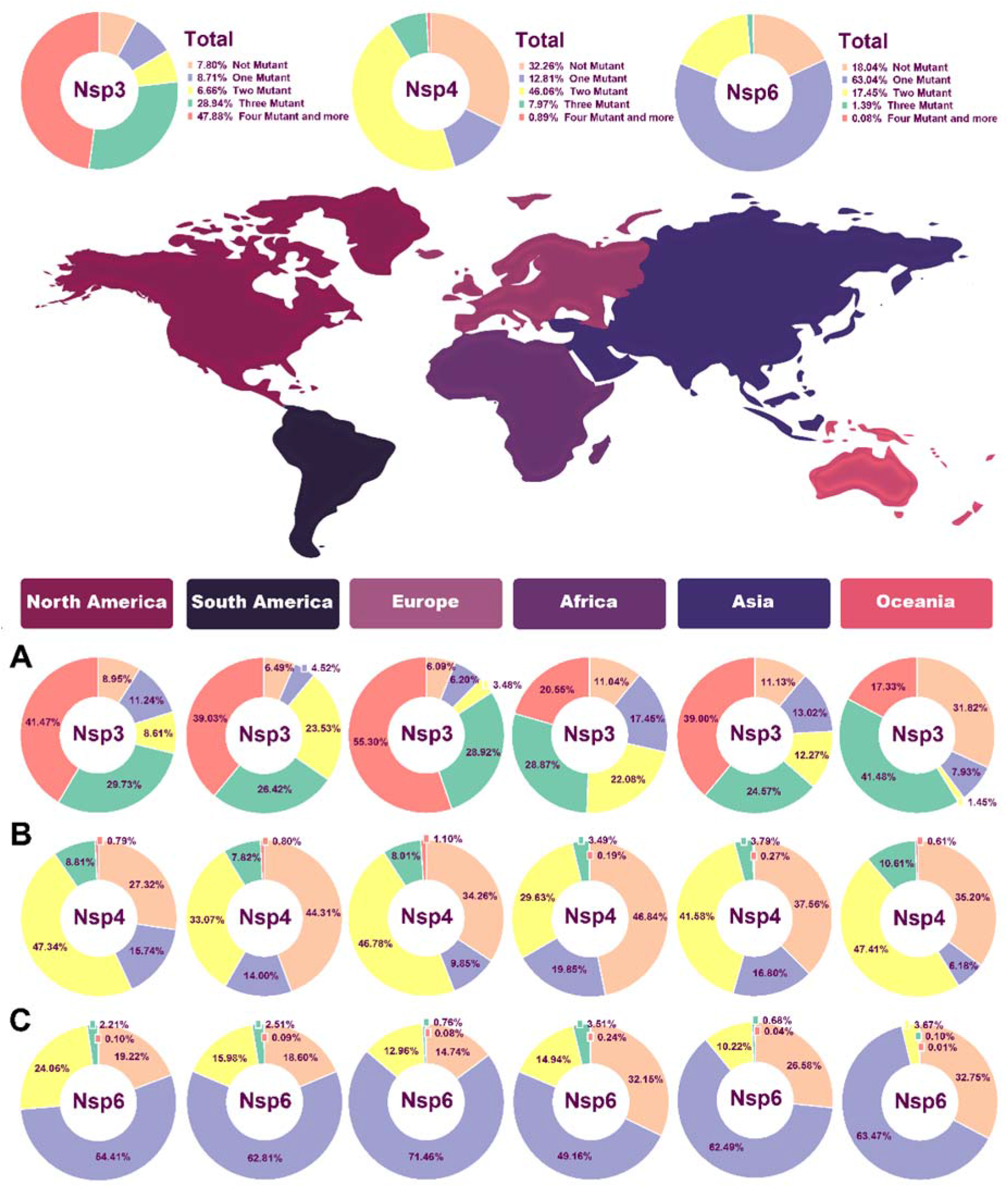
Pie chart plot of the number of mutations in NSPs of SARS-COV-2 as of January 2021. A) NSP3 B) NSP4 C) NSP6. The light orange, purple, yellow, green, and dark pink colors represent zero, one, two, three, and four or more mutations, respectively.

To follow the effective geographical zone on mutations pattern, mutations were categorized based on continents in table 2. Based on our results, North America NSP3 data represented no mutations in 8.94% of sequences, one mutation in 11.23%, two mutations in 8.6%, three mutations in 29.73% of AASs, and 41.47% of AASs showed more than four mutations in their AASs (Fig. 3A). North America NSP4 data demonstrated no mutations in 27.32% of sequences, one mutation in 15.74%, two mutations in 47.33%, three mutations in 8.8% of AASs, and 0.78% of AASs showed more than four mutations in their AASs (Fig. 3B). NSP6 data from North America represented no mutations in 19.22% of sequences, one mutation in 54.41%, two mutations in 24.05%, three mutations in 2.2% of AASs, and 0.09% of AASs showed more than four mutations in their AASs (Fig. 3C).

**Table 2.**
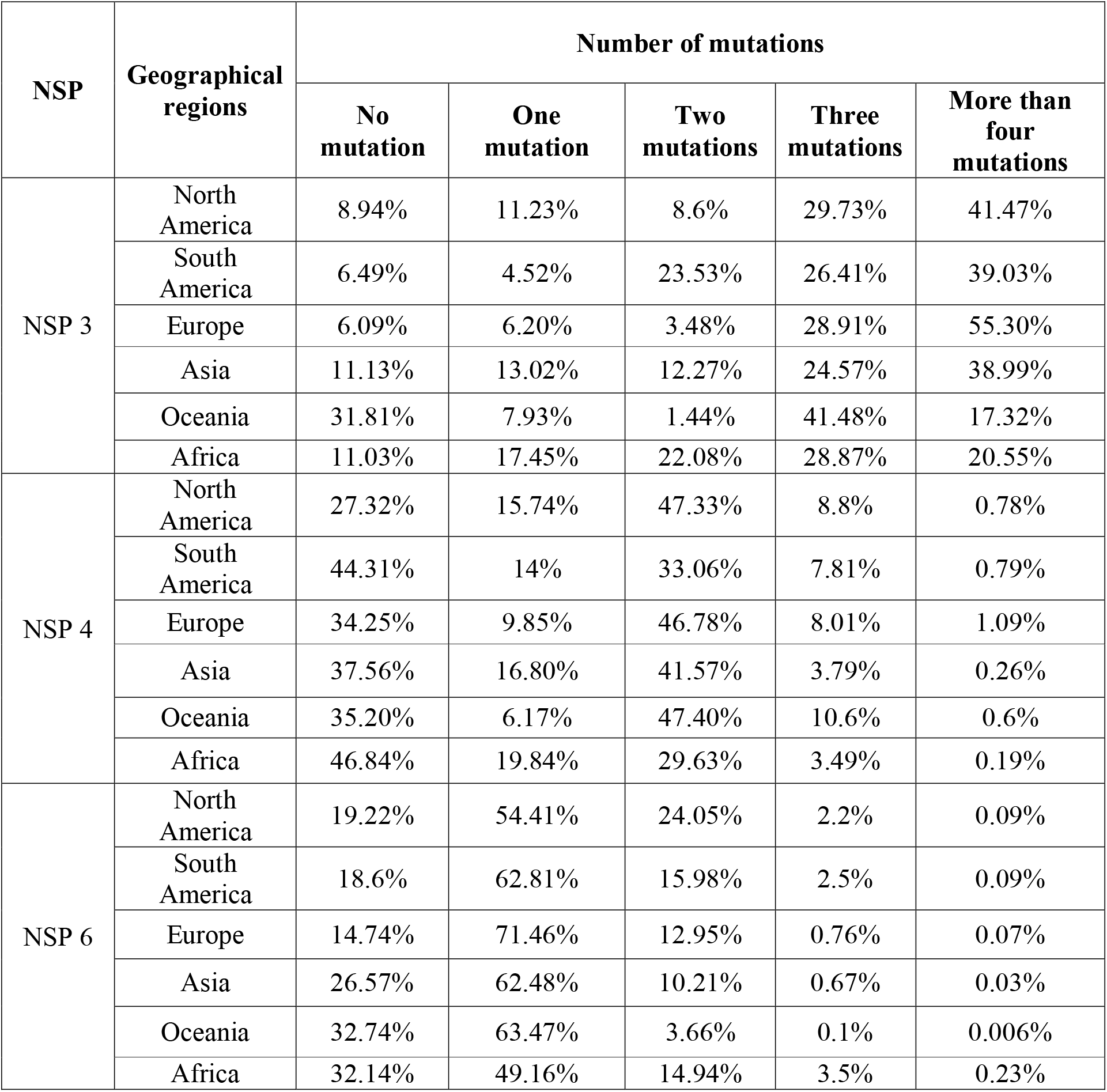
The number of NSP3, NSP4, and NSP6 mutations based on geographical regions

South America NSP3 data demonstrated no mutations in 6.49% of sequences, one mutation in 4.52%, two mutations in 23.53%, three mutations in 26.41% of AASs, and 39.03% of AASs showed more than four mutations in their AASs (Fig. 3A).South America NSP4 data represented no mutations in 44.31% of sequences, one mutation in 14%, two mutations in 33.06%, three mutations in 7.81% of AASs, and 0.79% of AASs showed more than four mutations in their AASs (Fig. 3B). NSP6 data from South America demonstrated no mutations in 18.6% of sequences, one mutation in 62.81%, two mutations in 15.98%, three mutations in 2.5% of AASs, and 0.09% of AASs showed more than four mutations in their AASs (Fig. 3C).

For NSP3 in Europe, no mutation was detected in 6.09% of sequences, one mutation was detected in 6.20%, two mutations in 3.48%, three mutations in 28.91% of AASs, and 55.30% of AASs showed more than four mutations in their AASs (Fig. 3A).For NSP4 in Europe, no mutation was detected in 34.25% of sequences, one mutation in 9.85%, two mutations in 46.78%, three mutations in 8.01% of AASs, and 1.09% of AASs showed more than four mutations in their AASs (Fig. 3B). Europe NSP4 data showed no mutation in 14.74% of sequences, one mutation in 71.46%, two mutations in 12.95%, three mutations in 0.76% of AASs, and 0.07% of AASs showed more than four mutations in their AASs (Fig. 3C).

Data regarding NSP3 in Asia demonstrated no mutation in 11.13% of sequences, one mutation in 13.02%, two mutations in 12.27%, three mutations in 24.57% of AASs, and 38.99% of AASs showed more than four mutations in their AASs (Fig. 3A). For NSP4 in Asia, no mutation was observed in 37.56% of sequences, one mutation in 16.80%, two mutations in 41.57%, three mutations in 3.79% of AASs, and 0.26% of AASs showed more than four mutations in their AASs (Fig. 3B). NSP6 data from Asia demonstrated no mutation in 26.57% of sequences, one mutation in 62.48%, two mutations in 10.21%, three mutations in 0.67% of AASs, and 0.07% of AASs showed more than four mutations in their AASs (Fig. 3C).

Oceania NSP3 data demonstrated no mutation in 31.81% of sequences, one mutation in 7.93%, two mutations in 1.44%, three mutations in 41.48%, and 17.32% AASs demonstrated more than four mutations in their AASs (Fig. 3A). For NSP4, Oceania data indicated no mutation in 35.20% of sequences, one mutation in 6.17%, two mutations in 47.40%, three mutations in 10.6% of AASs, and 0.6% AASs demonstrated more than four mutations in their AASs (Fig. 3B).NSP6 data from Oceania represented no mutation in 32.74% of sequences, one mutation in 63.47%, two mutations in 3.66%, three mutations in 0.1% of AASs, and 0.006% AASs demonstrated more than four mutations in their AASs (Fig. 3C).

Finally, NSP3 data related to Africa revealed no mutation in 11.03% of sequences, one mutation in 17.45%, two mutations in 22.08%, three mutations in 28.87%, and 20.55% of AASs represented more than four mutations in their AASs (Fig. 3A). For NSP4 data related to Africa, no mutation was observed in 46.84% of sequences, one mutation in 19.84%, two mutations in 29.63%, three mutations in 3.49% of AASs, and 0.19% of AASs represented more than four mutations in their AASs (Fig. 3B).NPS6 data from Africa revealed no mutation in 32.14% of sequences, one mutation in 49.16%, two mutations in 14.94%, three mutations in 3.5% of AASs, and 0.23% of AASs represented more than four mutations in their AASs (Fig. 3C).

Interestingly, in NSP3, Oceania had the lowest frequency of more than four mutations group with 17.32%. In NSP4, the highest and the lowest percentages of two and three mutations groups with 47.40% and 10.6% were also related to Oceania.

For NSP6, in all continents except Africa more than half of samples had two mutations including 71.46%, 63.47%, 62.81%,62.48% and 54.41% in Europe, Oceania, South America, Asia and North America, respectively.

Since at least one mutation has occurred in most NSP3, NSP4, and NSP6 sequences in all geographical areas, our data indicate that NSP3, NSP4, and NSP6 are critical points of the mutation. Counting the number of mutations in AASs is not sufficient to anticipate the impact of every single mutation, as some changes can occur repeatedly. However, others can occur in just a few samples. For this purpose, a heat map was prepared to study the frequency of mutations on every section of sequences of NSP3, NSP4, and NSP6 proteins.

Based on total mutation outcomes, the results from NSP3 demonstrated that the regions including the highest mutations were 1358 to 1552 (0.41%), 1164 to 1358 (0.34%), 388 to 582 (0.32%), and 776 to 970 (0.17%), respectively (Fig. 4A). For NSP4, the hotspot zones were identified in the 150 to 200 region (1.01%) and after that in the positions of 450 to 500 (1.11%), 400 to 450 (0.15%) (Fig.4B). Regarding NSP6, the highest mutations frequency occurred in the position of 58 to 87 (2.43%) and then, in the positions of 145 to 174 (0.42%) and 174 to 203 AA (0.31%) (Fig. 4C). The complete list of total and per continents mutations, their frequencies and percentages is brought in the Supplementory file 1 A to C.

**Fig. 4.**
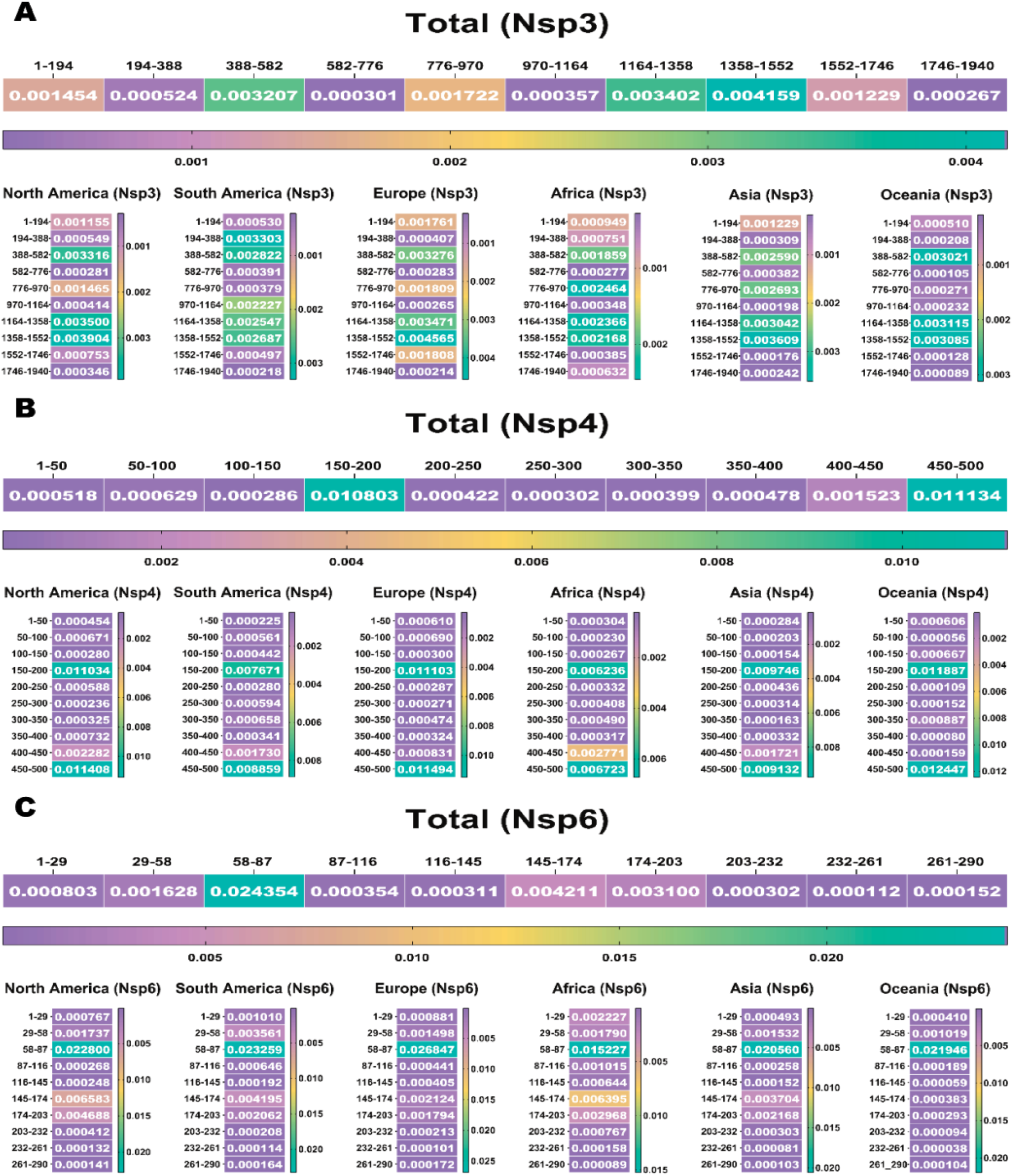
The heat maps indicate the rate of mutations per AA based on geographical zones and show the highest frequencies. A) indicates the rate of mutation, per 200 amino acids of the NSP-3 protein. The highest frequency rate occurred in the 1358 to 1552 amino acid sequence of the NSP-3 protein. B) indicates the rate of mutation per 50 amino acids of the NSP-4 protein. The highest frequency rates occurred in the 150 to 200 and 450 to 500 amino acid sequence of the NSP-4 protein. C) indicates the rate of mutation per 30 amino acids of the NSP-6 protein. The highest frequency rate occurred in the 58 to 87 amino acid sequence of the NSP-6 protein.

Based on the continents, for NSP3, in Asia, Europe and North America the position of 1358 to 1552, and in Oceania, Africa and South America the areas of 1164 to 1358, 776 to 970 and 194 to 388, respectively, were the most hotspot regions. For NSP4 in Africa, Europe, Oceania, North America and South America the position of 450 to 500, and in Asia the area of 150 to 200 were the most hotspot regions. For NSP6 in all continents the position of 58 to 87 was the most hotspot region. The complete list of total and per continents mutations, their frequencies and percentages is noted in the supplementory file 2 A to C

### 3.2 Common mutations in NSP 3, 4, and 6

To investigate sustained mutations of NSP3, NSP4, and NSP6 the position of mutations in the protein structure and their frequency were investigated. Table 3 shows common mutations in NSP 3, 4, and 6 based on two factors: The presence of all mutations in all continents and having a frequency above 0.2 (20%).

**Table 3.** Common NSP 3, 4, and 6 point mutations globally which present among all continents and frequency > 0.2

| Rank | Residue | Percentage% |
| --- | --- | --- |
| NSP3 1<sup>st</sup> | P(1228)L | 54.48% |
| NSP3 2<sup>nd</sup> | P(1469)S | 54.41% |
| NSP3 3<sup>rd</sup> | A(488)S | 53.86% |
| **NSP4 1<sup>st</sup>** | T(492)I | 54.84% |
| **NSP4 2<sup>nd</sup>** | V(167)L | 52.81% |
| **NSP6** | T(77)A | 69.85% |

According to our results, the most frequent mutation for NSP3 until November 2021 was P1228L (0.544826) (Fig. 5A) and T492I (0.54842) for NSP4 at the same time (Fig. 5B). For NSP6, T77A (0.69856) (Fig. 5C) was the most prevalent until November 2021.

**Fig. 5.**
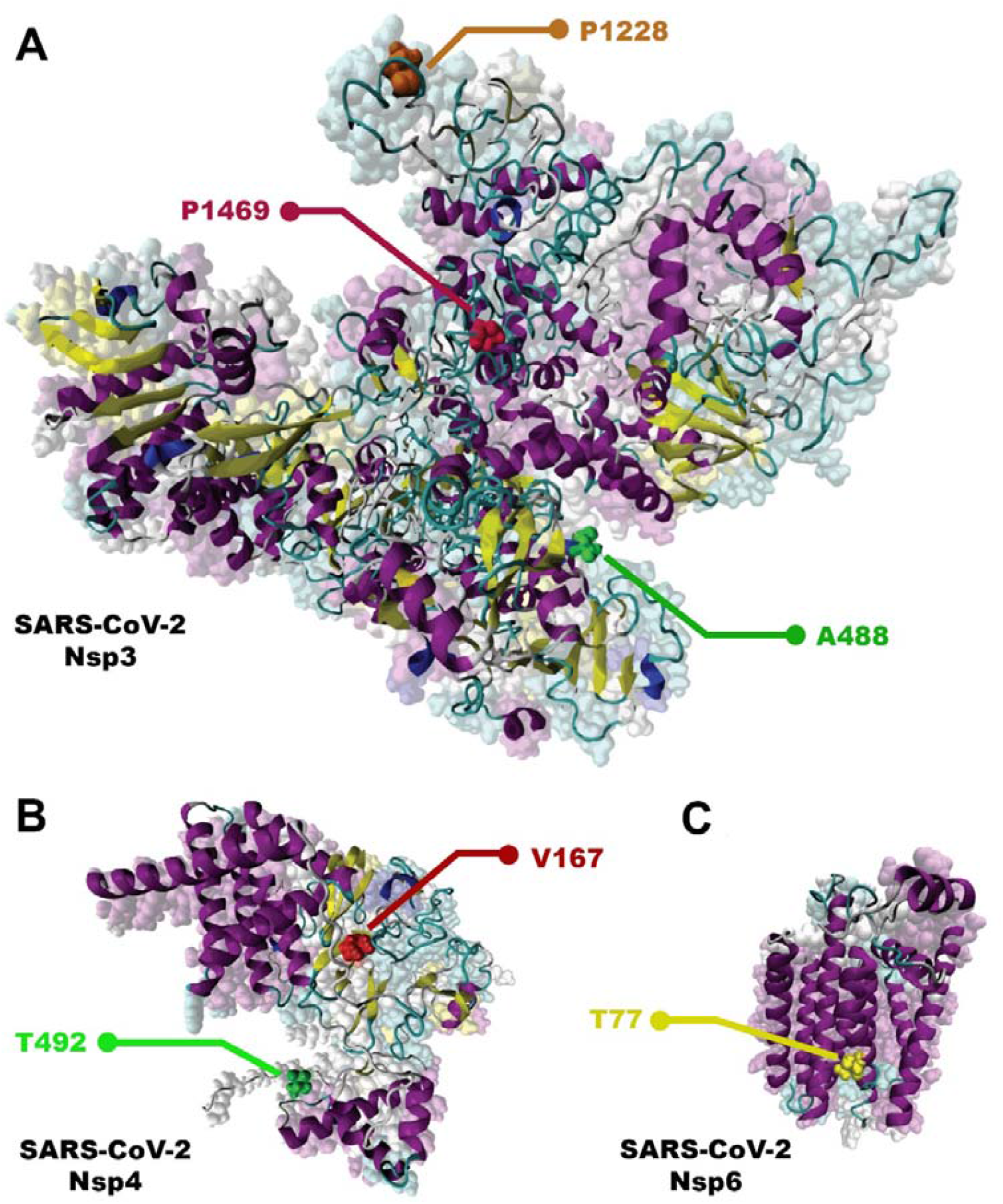
Most frequence NSP3, NSP4, and NSP6 mutations. A) NSP 3; P1228L, P1469 and A488. B) NSP 4; T492I and V167L. C) NSP 6; T77A.

### 3.3 Statistical occurrence of NSP 3, 4, and 6 mutations by continents

Based on the continents, the statistical occurrence of these top frequent mutations of NSP 3, 4, and 6 are listed in Table 4, 5, and 6, according to the presence of all mutations in all continents and having the frequency above 0.2.

**Table 4.** The incidence of the global NSP3 mutations based on their continental distribution

| Residue | Variant Percentage |  |  |  |  |  |
| --- | --- | --- | --- | --- | --- | --- |
|  | North America | South America | Europe | Asia | Oceania | Africa |
| P(1228)L | 53.87% | 39.48% | 57.23% | 49.60% | 56.35% | 26.71% |
| P(1469)S | 53.72% | 45.17% | 57.19% | 48.14% | 56.32% | 27.26% |
| A(488)S | 53.74% | 39.23% | 57.16% | 42.93% | 56.14% | 26.03% |

**Table 5.** The incidence of the global NSP4 mutations is based on their continental distribution

| Residue | Variant Percentage |  |  |  |  |  |
| --- | --- | --- | --- | --- | --- | --- |
|  | North America | South America | Europe | Asia | Oceania | Africa |
| T(492)I | 56.00% | 43.56% | 56.74% | 45.26% | 61.01% | 31.78% |
| V(167)L | 53.51% % | 34.18% | 54.71% | 48.18% | 58.11% | 28.14% |

**Table 6.** The incidence of the global NSP6 mutations based on their continental distribution

| Residue | Variant Percentage |  |  |  |  |  |
| --- | --- | --- | --- | --- | --- | --- |
|  | North America | South America | Europe | Asia | Oceania | Africa |
| T(77)A | 65.55% | 66.90% | 77.09% | 58.26% | 62.75% | 40.49% |

Interestingly, for NSP3, P1228L, P1469S, and A446S, for NSP 4, T492I, and V167L, and NSP 6, T77A mutations were present among all continents as the top three mutations, and their frequencies were more than 0.2. The P1469S mutation was found as the most prevalent mutation in South America and Africa, the second most prevalent in Europe, Asia, Oceania, and the third most prevalent in South America. A446S mutation has been observed as the second most prevalent mutation in North America and the third most prevalent mutation in Africa, Asia, Europe, South America, and Oceania (Fig. 6A). T492I and V167L in NSP 4 were present in all continents as the mutation with the highest prevalence rate (Fig. 6B). The T77A mutation was present among the continents as a top mutation (Fig. 6C). The complete list of total and continents mutations and their frequencies is noted in the supplementory file 3 A to C.

**Fig. 6.**
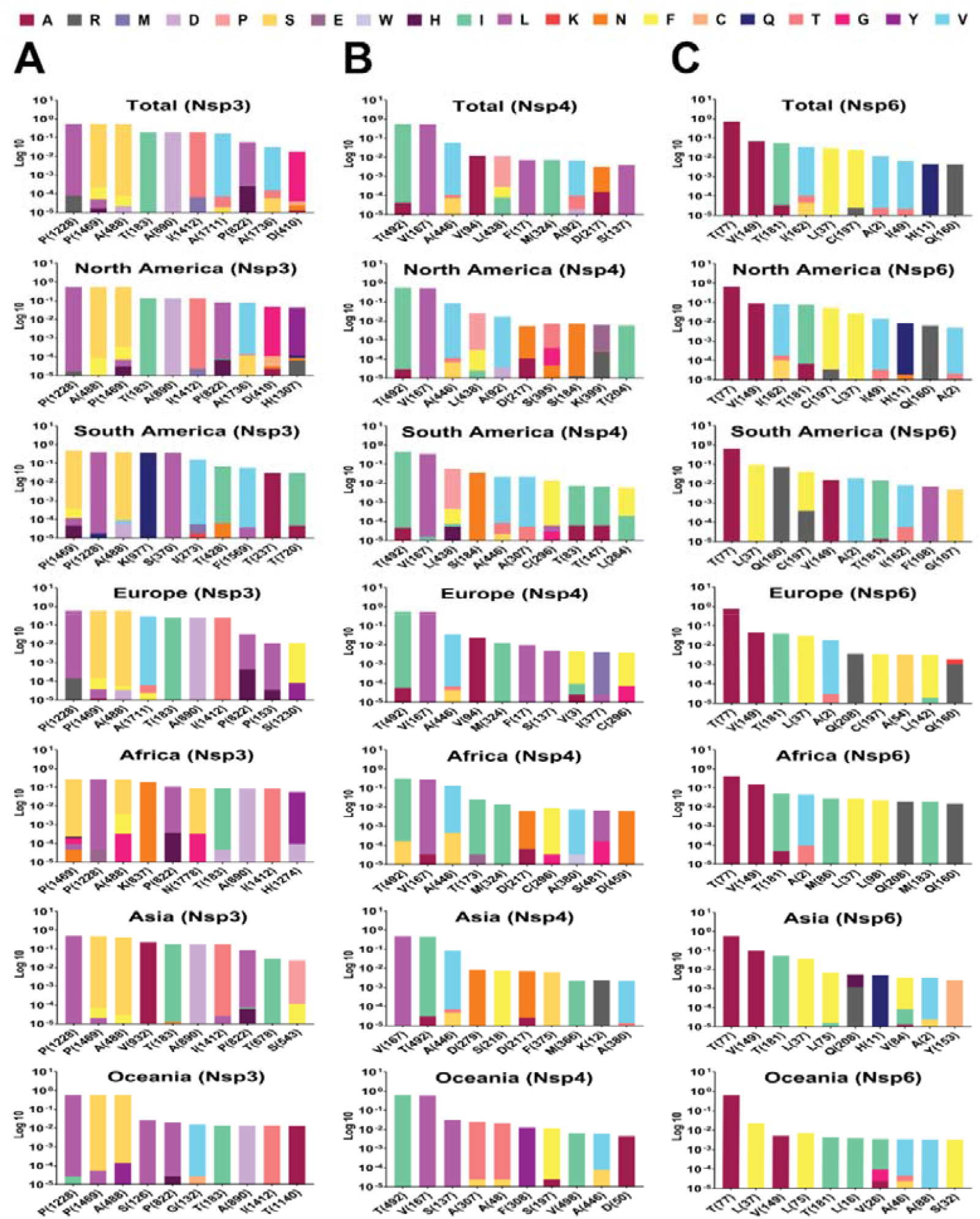
Top 10 mutations in (A) NSP 3, (B) 4 and (C) 6, with the highest frequency worldwide and geographic areas including North America, South America, Europe, Asia, Oceania, and Africa. The position of altered amino acids and substituted ones is shown differently based on the frequency percentage of substituted AA. The mutation frequency was estimated for each of them by normalizing the number of genomes carrying a given mutation in a desired geographic areas.

### 3.4 NSP3, 4, and 6 mutations Natrual selections

To determine the appearance of each mutation, we analyzed AASs from each geographic area over time by classifying them according to the month of sample collection from November 01, 2019, till November 01, 2021, as indicated in the GISAID (www.gisaid.org) [35-37]. The detailed distribution of the top ten high mutation rates of NSP3, 4, and 6 variants from the world and each continent is provided by the month of sample collection and illustrated in Fig.6. Next, we investigated the continuation of mutations with the prevalence rate above 0.2 per AASs collected every month.

For NSP3, P1228L, P1469S, and A488S mutations emerged at the pandemic’s beginning. Interestingly, their prevalence has risen significantly after 31th of May 2021 in all geographical regions (Fig. 7A). For NSP4, T492I mutation appeared at the beginning of the pandemic. Its prevalence has increased significantly from May 2021 in all geographical regions except Asia. V167L mutation was as same as T492I, but the mutation trend reduced in the last month (Fig.7B). For NSP6, T77A mutation began to emerge at the beginning of the pandemic. In most geographical regions, its prevalence has risen sharply from May 2021 (Fig.7C). The complete list of total and per continent mutations and their frequencies is noted in the supplementory file 4 A to C.

**Fig. 7.**
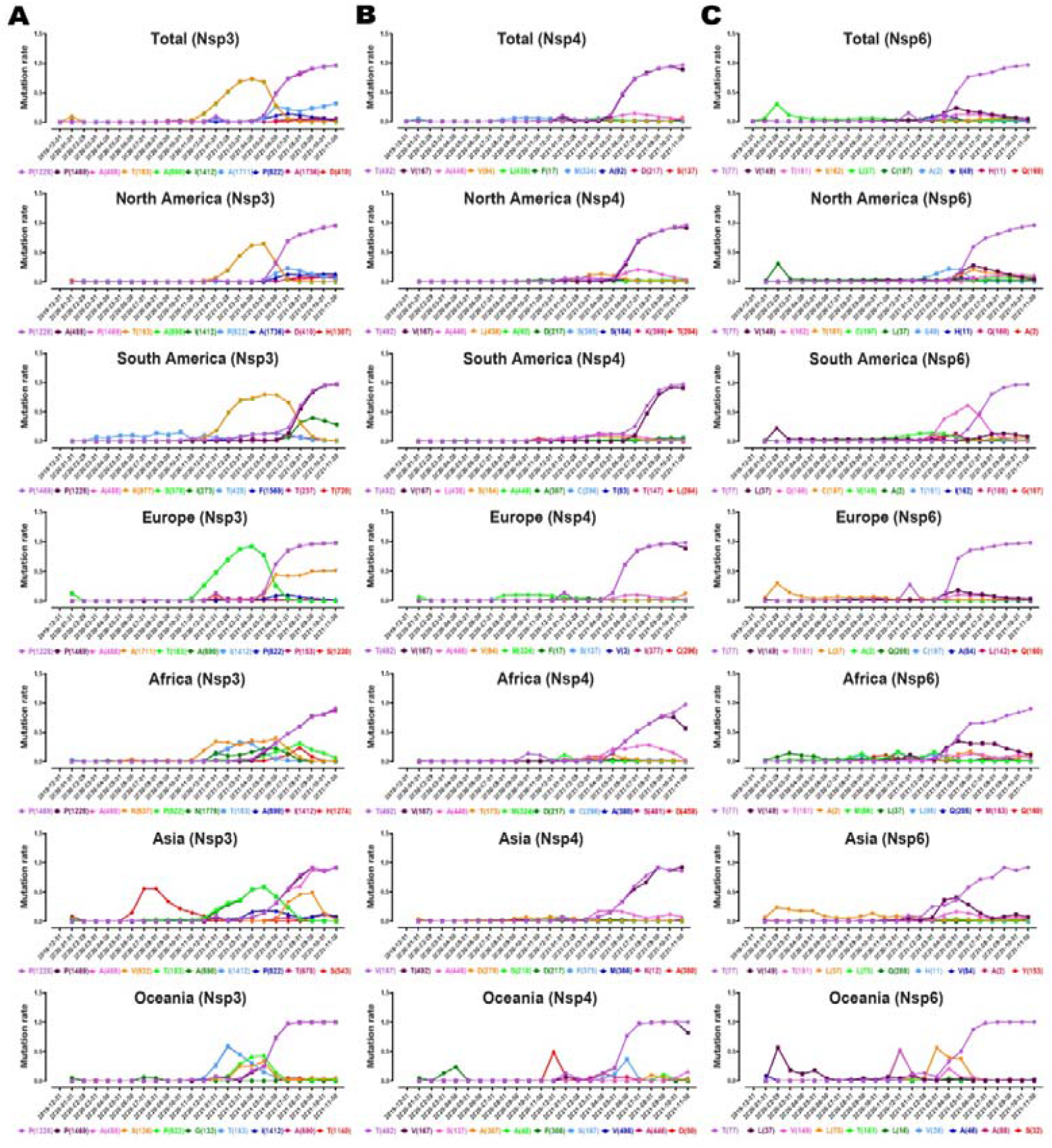
Plots time evolution trajectories of top ten high-rate mutations of (A) NSP 3, (B) 4, and (C) 6, worldwide and different geographic areas including North America, South America, Europe, Asia, Oceania, and Africa. According to the month of sample collection, data is computed as the number of AASs having a given mutation over the total number of AASs according to the month of sample collection.

### 3.5 NSP3, NSP4, and NSP6 corelations and Epistasis

Interestingly, the regressions between NSP3 and NSP4 (Fig.8A), NSP3 and NSP6 (Fig.8B), and NSP4 and NSP6 (Fig.8C) showed noticeable correlations.

**Fig. 8.**
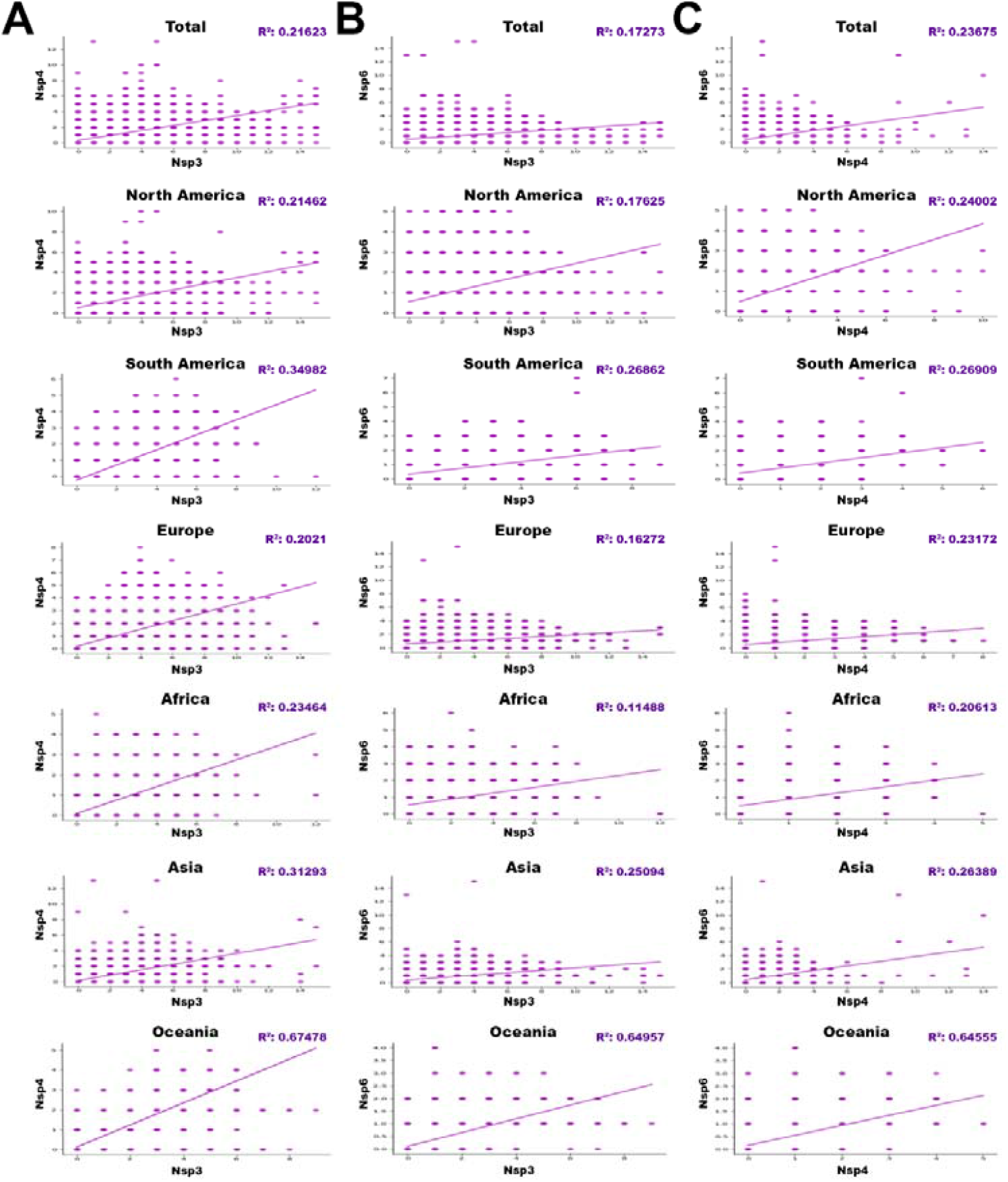
The regressions between (A)NSP3 and NSP4, (B)NSP3 and NSP6, and (C) NSP4 and NSP6 with p-value<0/05, based on total and different geographic areas including North America, South America, Europe, Asia, Oceania, and Africa. (r “parameter is directly show correlation and Epistasis between genes.)

Regressions with p-value<0.05, are indicated in the table 7 based on total in different worldwide zones (Fig.8).

**Table 7.** Regressions with p-value<0.05 between NSP3 and NSP4, NSP3 and NSP6, and NSP4 and NSP6. “r “parameter directly shows correlation and Epistasis between candidate genes.

| Geographical regions | r Value |  |  |
| --- | --- | --- | --- |
|  | NSP3-NSP4 | NSP3-NSP6 | NSP4-NSP6 |
| Total | 0.4650 | 0.41560 | 0.48656 |
| Africa | 0.48439 | 0.33894 | 0.45401 |
| Asia | 0.55940 | 0.50093 | 0.51370 |
| Europe | 0.44955 | 0.40338 | 0.48137 |
| North America | 0.46326 | 0.41982 | 0.48991 |
| Oceania | 0.82145 | 0.80595 | 0.80346 |
| South America | 0.59145 | 0.51828 | 0.51873 |

The complete list of total and per continent regressions and their frequencies is noted in the supplementory file 5 A to C.

## 4. Discussion

Coronavirus 2019 (COVID-19) epidemy due to the SARS-CoV-2 virus has forced the scientific community to give a strong response by performing massive studies, research and innovations around the virus, then controlling its spreading and threatening effects. In fact, the key stage of the SARS-CoV-2 life cycle is virus replication in infected cells. During this process, several viral proteins facilitate genome replication. Some of these proteins are related to non-structural proteins (NSP), like that NSP3, NSP4, and NSP6. Due to their important role in virus replication and pathogenicity, we investigated the occurrence of mutations in their hot spots regions from January 2020 to January 2022 with about 6.5 million AASs of SARS-CoV2 from GISAID [35-37]. Then we selected the key mutations of each protein according to their presence in all studied continents with the highest mutations frequency then, evaluated the correlation between proteins. Next, we examined the amino acids changes in mutations by their degree of hydrophobicity (HΦ), using previous studies [38] (Table 9), and predicted the effect of mutations on the stability and replication of NSP3, NSP4, and NSP6 in virus genome. We also obtained the main target genes of proteins in the host cell using the Human Protein Atlas Databasis (https://www.proteinatlas.org/humanproteome/sars-cov-2) and based on them we completed our predictions.

**Table 9.** The degree of hydrophobicity of amino acids

| AA | $H\Phi$ |
| --- | --- |
| Phe | 20/000 |
| Met | 17/620 |
| Ile | 16/667 |
| Leu | 16/190 |
| Val | 14/762 |
| Pro | 13/810 |
| Cys | 12/857 |
| Ala | 10/952 |
| Trp | 8/095 |
| Gly | 7/143 |
| Lys | 6/350 |
| Tyr | 6/259 |
| Thr | 3/810 |
| Ser | 3/129 |
| His | 2/064 |
| Gln | 1/723 |
| Asn | 1/451 |
| Glu | 1/400 |
| Asp | 1/166 |
| Arg | 0/79 |

Data analysis showed the P1228L, P1469S, and A488S as NSP3 mutations. In P1228L mutation (proline to leucine at AA., 1228), proline with HΦ∼13.5 converts to leucine with HΦ∼16.0. As regards, increasing the degree of hydrophobicity enhanced the tendency of water to move non-polar groups of molecules away from themselves and join them together in the aqueous environment of the cytoplasm, reducing the stability of the AA [39]. The P1228L is located in the α–helical region of NSP3 and it is placed in the cell cytoplasm. Therefore, leucine may reduce the stability of NSP3 in the P1228L location.

In P1469S mutation (proline to serine at AA. 1469), proline with HΦ∼13.5 changs to serine with HΦ∼3.0. Serine with law hydrophobicity known as a hydrophilic AA. P1469S is located at the T.M. domain of NSP3 and situated in the cell membrane, where the tendency to absorb fat is more than water [40]. Thus, it might decline the stability of NSP3 in P1469S place. Also, in another study, Kirchdoerfer, R.N et al. have found that mutations P1228L and P1469S are likely to reduce protein stability, but, since these mutations are not located in the protease domain (aa 783–1036), it is still unclear whether these mutations negatively affect the performance of the protease [41]. Based on previous findings and studies, we predicted that the P1228L and P1469S mutations would be reduced their positional stability in NSP3. These mutations were placed near the protein sequence terminal, and this area has been important in connecting NSP3 to NSP4, so we hypothesized that they might have a negative effect on bindings.

In contrast, A488S mutation (alanine to serine at AA. 488), alanine with HΦ∼11.0 transforms to serine with HΦ∼3.0 in the cytoplasm. Conversion of hydrophobic alanine to hydrophilic serine probably increase NSP3 stability in that area, on the other, this mutation set in SUD domain of the protein that might exhibit a binding selectivity towards G-quadruplex (G4). This feature may contribute to the characterization of their role in forming the replication/transcription viral complex (RTC). According to this information, we predicted that the A488S might increase the stability of NSP3 in its area and amplify the connection of SUD to G4s, thereby having a positive effect on RTC.

STAT1, PARG-345, PARG-329, PARP9 genes could be known as suitable targets for NSP3 in the host cells. Since the STAT1 gene has been identified as an activator of the transcription complex [42], it may play a role in enhancing the transcription of the protein.At the whole, our meta AAs sequence data analysis study have identified the mutations T492I and V167L in NSP4. In T492I mutation (threonine to isoleucine at AA 492), threonine with HΦ∼6.0 transforms to isoleucine with HΦ∼16.5, and in V167L (Valine to Leucine at AA 167) Valine with HΦ∼14.5 changs to Leucine with HΦ∼16.0. Thus, the mutations are associated with an increase in the degree of hydrophobicity. Both T492I and V167L are located in M domain of NSP4, known as the cell’s bilayer membrane. This domain is also involved in forming double-membrane vesicles (DMV) and copies of virus fragments associated with RTC replication. By the evidence, we predicted that these mutations might increase the stability of NSP4 in their region and enhance the RTC. For NSP4, ALG11, DNAJC11, IDE, NUP210, TIMM10, TIMM10B, TIMM29, TIMM9 genes presented as targets in host cells. Among these, DNAJC11, TIMM10, and TIMM9 act as chaperone by binding to the target protein [43-45]. Thus, it may facilitate folding and prevent its decomposition.

In the current study, the T77A mutation was investigated in NSP6. In this mutation (threonine to alanine at AA 77), threonine with HΦ∼6.0 turns to alanine with HΦ∼11.0, which increases hydrophobicity. T77A is located in TM domain of NSP6, which is associated with the cell membrane. Also, the NSP6, ATP13A3, ATP5MG, ATP6AP1, and SIGMAR1 genes are shown as targets in host cells. ATP13A3 acts as an ATP pump to transport cations across the membrane and increase its stability [46]. Since the function of NSP6 was to induce autophagy in favor of viral infection, we predicted that the mutation would be effective on the stability of NSP6 in that situation and possibly have a positive effect on virus infectivity.

According to our data, the Oceania is known as a highly epistatic continent among anothers. Therefore, for the first time, we reported that the NSP3, NSP4, and NSP6 mutations had an significant epistatic correlation (P<0.05). We also believe that the mutations of these proteins are probably related to each other in terms of molecular function.

Finally, we suggest for performing molecular dynamic protein modeling for our candidate mutations to predict molecular functions and structural variations and then, *in vitro* evaluation of the findings.

## Supporting information

Additionalfile1A, NSP3.xlsx

Additionalfile1B, NSP4.xlsx

Additionalfile1C, NSP6.xlsx

Additionalfile2A, NSP3.xlsx

Additionalfile2B, NSP4.xlsx

Additionalfile2C, NSP6.xlsx

Additionalfile3A, NSP3.xlsx

Additionalfile3B, NSP4.xlsx

Additionalfile3C, NSP6.xlsx

Additionalfile4A, NSP3.xlsx

Additionalfile4B, NSP4.xlsx

Additionalfile4C, NSP6.xlsx

Additionalfile5A, NSP3-4

Additionalfile5B, NSP3-6

Additionalfile5C, NSP6-4

## Data Availability

All data were retrieved from January 2020 until January 2022 from GISAID (www.gisaid.org) Database through coordination with Erasmus Medical Center

## Declaration of competing interest

The authors declare that they have no conflicts of interest that might be relevant to the contents of this manuscript and the research was carried out regardless of commercial or financial relationships that may cause any conflict of interests.

## Data availability

The raw data supporting the conclusions of this article is available in supplementary file(s).

## Supplementary

Supplemental data associated with the current study have been gathered. Click to download.

- Additionalfile1A, NSP3.xlsx
- Additionalfile1B, NSP3.xlsx
- Additionalfile1C, NSP3.xlsx
- Additionalfile2A, NSP3.xlsx
- Additionalfile2B, NSP3.xlsx
- Additionalfile2C, NSP3.xlsx
- Additionalfile3A, NSP3.xlsx
- Additionalfile3B, NSP3.xlsx
- Additionalfile3C, NSP3.xlsx
- Additionalfile4A, NSP3.xlsx
- Additionalfile4B, NSP3.xlsx
- Additionalfile4C, NSP3.xlsx
- Additionalfile5A, NSP3-4
- Additionalfile5B, NSP3-4
- Additionalfile5C, NSP3-4

## Authors’ contributions

<u>K.R, M.M, M.M.S, S.T, B.M.</u> contributed to data collection, M.G, M.R.G, <u>K.R, H.F. contribute to study design, K.R, M.M design workflow and code, data analysis. K.R, M.M, B.M data visualization. H.f, M.S, M.A, S.T wrote the manuscript. M.M, K.R, B.M</u> monitored the accuracy of Additional data. B.M designed graphical contents. M.G, M.R.G <u>have edited and supervised the work.</u>

## Acknowledgments

The authors thank all of the researchers who have shared genome data openly via the Global Initiative on Sharing All Influenza Data (GISAID)

## Key point

- The most frequent mutation until November 2021 for NSP3, NSP 4 and NSP 6 was P1228L, T492I and T77A respectively. Also the most frequent mutation rate, belonged to T77A in NSP 6 with 0.69856 frequency.
- The highest rate of the global mutations based on their continental distribution for P1228L of NSP3, T492I of NSP4 and T77A of NSP6, belonged to Europe.
- Totally, the most conserved area for NSP3, NSP4 and NSP6 was, 1746_1940, 150_100and 232_261 AA.
- The regressions with p-value<0.05 between NSP3 and NSP4, NSP3 and NSP6, and NSP4 and NSP6 showed notable correlations especially in the Oceania.
- We predicted the effect of mutations on the stability and replication of NSP3, NSP4 and NSP6 in virus genome.

## Notes

### Competing Interest Statement

The authors have declared no competing interest.

### Funding Statement

No funded

